# Changes in body and interoceptive awareness after Cognitive Multisensory Rehabilitation or Qigong in adults with spinal cord injury

**DOI:** 10.64898/2025.12.12.25342154

**Authors:** Sydney Carpentier, Ann Van de Winckel

**Author notes:** Corresponding author: Sydney Carpentier.

## Abstract

Brain imaging studies have demonstrated that adults with spinal cord injury (SCI) exhibit deficits in body and interoceptive awareness. However, there is limited research on the degree and impact of these deficits. Few clinical trials have examined interventions to improve body and interoceptive awareness in this population.

We compared scores on the Multidimensional Assessment of Interoceptive Awareness, Version 2 (MAIA-2) and the Revised Body Awareness Rating Questionnaire (BARQ-R) between adults with SCI and uninjured adults. We also assessed changes in body and interoceptive awareness in adults with SCI following one of two body awareness interventions, i.e., Qigong and Cognitive Multisensory Rehabilitation (CMR).

Adults with SCI reported worse interoceptive awareness than uninjured adults on the MAIA-2 dimensions “Not-distracting” [Median(IQR), SCI vs healthy=1.33 (1.17) vs 2.30 (1.5); *p*<.0001] and “Trust” [3.50 (1) vs 3.70 (1.3); *p*=.02]. Conversely, they scored better on “Noticing” [3.50 (1.38) vs 3.00 (1.50); *p*=.035], “Attention Regulation” [3.50 (1) vs 2.70 (1.3); *p*<.0001], “Self-regulation” (3.75 (1) vs 3.00 (1.5); *p*<.0001], and “Body Listening” [3.75 (1) vs 2.30 (1.70); *p*<.0001]. Adults with SCI displayed worse body awareness, indicated by higher BARQ-R scores [18 (6.50) vs 15 (7); *p*<.0001].

Post-intervention results revealed improvements on the MAIA-2 dimension “Not-Worrying” [pre: 3.00 (0.80) vs post: 3.40 (1.40); *p*=.03] and the BARQ-R [pre: 18 (7) vs post: 16 (7); *p=*0.0004].

Given the demonstrated deficit in body and interoceptive awareness and the potential for improvement with interventions, our results encourage further exploration on how improving body and interoceptive awareness can impact quality of daily life.

## Introduction

Spinal cord injury (SCI) disrupts the sensory and motor information flow between the brain and the body, affecting impact mental body representations (MBR), which include body awareness and interoception.^1–5^ Whole-body awareness arises from integrating multisensory information,^3,5^ leading to an awareness of physical tension in the body and the positioning of body parts relative to one another. This information contributes to our awareness of the body in space.^4,6^ Interoception, a vital aspect of body awareness, pertains to the ability to perceive internal body signals.^6^

Brain imaging studies indicate that neural connections between regions, essential for sensorimotor function and MBR, are weaker in adults with SCI compared to healthy individuals.^7^ However, there is limited research on the extent and perception of these deficits in daily life for those with SCI. Additionally, few clinical trials focus on interventions aimed at enhancing body and interoceptive awareness in this population.^3,4^

Therefore, our two study objectives were: (1) to evaluate differences in body and interoceptive awareness between adults with SCI and uninjured adults, and (2) to assess changes in body and interoceptive awareness following interventions known to enhance body awareness, namely Qigong and a physical therapy approach called Cognitive Multisensory Rehabilitation (CMR).

## Method

### Data collection

Baseline data for adults with SCI were collected as a secondary analysis from 5 clinical trials (IRB#00011997; IRB#00022415; IRB#00014710; IRB#00018287; IRB#00008476) and 1 observational study (IRB#00018745). Uninjured adults completed the MAIA-2 and the BARQ-R at the Minnesota state fair (IRB#00005849) or as part of a control group in a clinical trial at the University of Minnesota (IRB#0008476).

Adults with SCI completed the Multidimensional Assessment of Interoceptive Awareness, Version 2 (MAIA-2) and the Revised Body Awareness Rating Questionnaire (BARQ-R) before and after either receiving remotely delivered Qigong (41min online video, 3x/week, for 12 weeks) IRB#00011997; IRB#00022415)^3–5^ or CMR (45min/session, 3x/week, for either 6 weeks (IRB#00008476)^7,8^ or 8 weeks in-person (IRB#00014710), or 12 weeks remotely over Zoom with a CMR-certified physical therapist (IRB#00018287).

### Outcome measures

MAIA-2 is a self-report measure encompassing 37 items that assess various aspects of interoceptive awareness, with scores ranging from 0 (never) to 5 (always). ^6^ Average scores are calculated for each dimension: **Noticing:** acknowledging sensations within the body; **Not-distracting**: no distraction from pain or discomfort; **Not-worrying**: not ruminating over pain or discomfort; **Attention Regulation:** sustaining focus on internal body sensations; **Emotional Awareness:** awareness of interactions between bodily sensations and emotions; **Self-regulation:** stress regulation through bodily awareness**; Body Listening**: addressing needs of the body; and **Trust:** whether one’s body feels like home and is experienced as safe.^6^ *Higher* scores indicate *better* interoceptive awareness.^6^

The BARQ-R, a self-report measure designed for adults with chronic musculoskeletal pain,^9^ consists of 12 items measuring awareness of bodily sensations and physical tension with scores ranging from 0 (completely disagree) to 3 (completely agree). The total sum score is calculated, with *lower* scores indicating *higher* body awareness.^9^

### Statistical Analyses

Demographic data were reported with descriptive statistics. The Mann-Whitney U test was employed to analyze between-group differences in MAIA-2 and BARQ-R scores, while the Wilcoxon Signed-Rank test was used to assess within-group changes pre-post-interventions. Data were reported as median and interquartile ranges. Statistical analyses were calculated with JMP statistical software version 18.

## Results

We enrolled 113 adults with SCI (48 women, 65 men), with an average age of 51±15 years and 13±12 years post-SCI. Of this cohort, at baseline 81 completed the MAIA-2 and 113 completed the BARQ-R. Those who practiced Qigong or received CMR and completed the MAIA-2 (n=43) and the BARQ-R (n=69), were demographically similar to the overall sample.

We recruited 471 uninjured community-dwelling adults (309 women, 161 men, 1 agender), with an average age of 50±18 years, completing a one-time MAIA-2 (n=439) and BARQ-R (n=471) assessment. **Table 1** displays additional demographic and descriptive information of both groups.

**Table 1.**
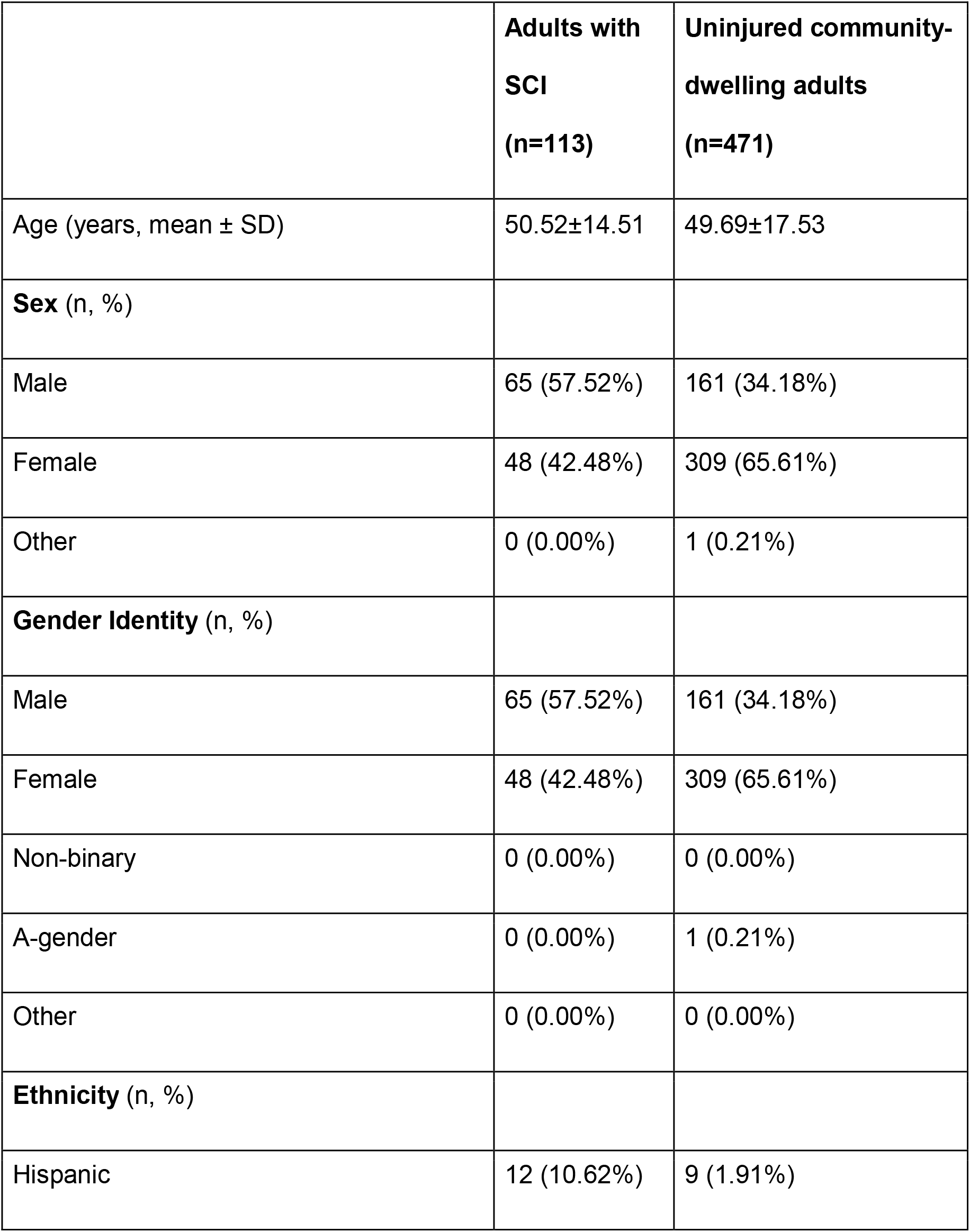
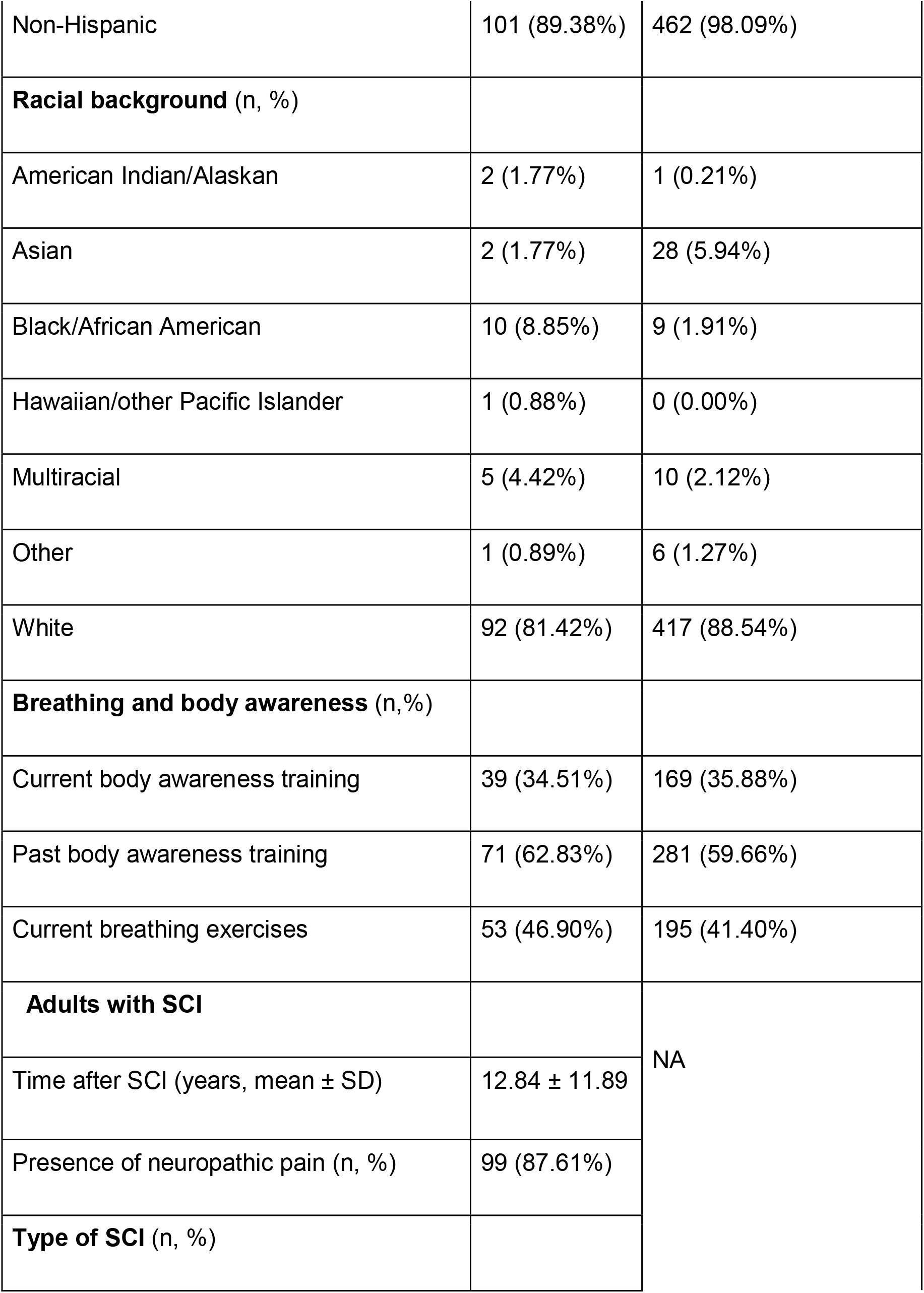
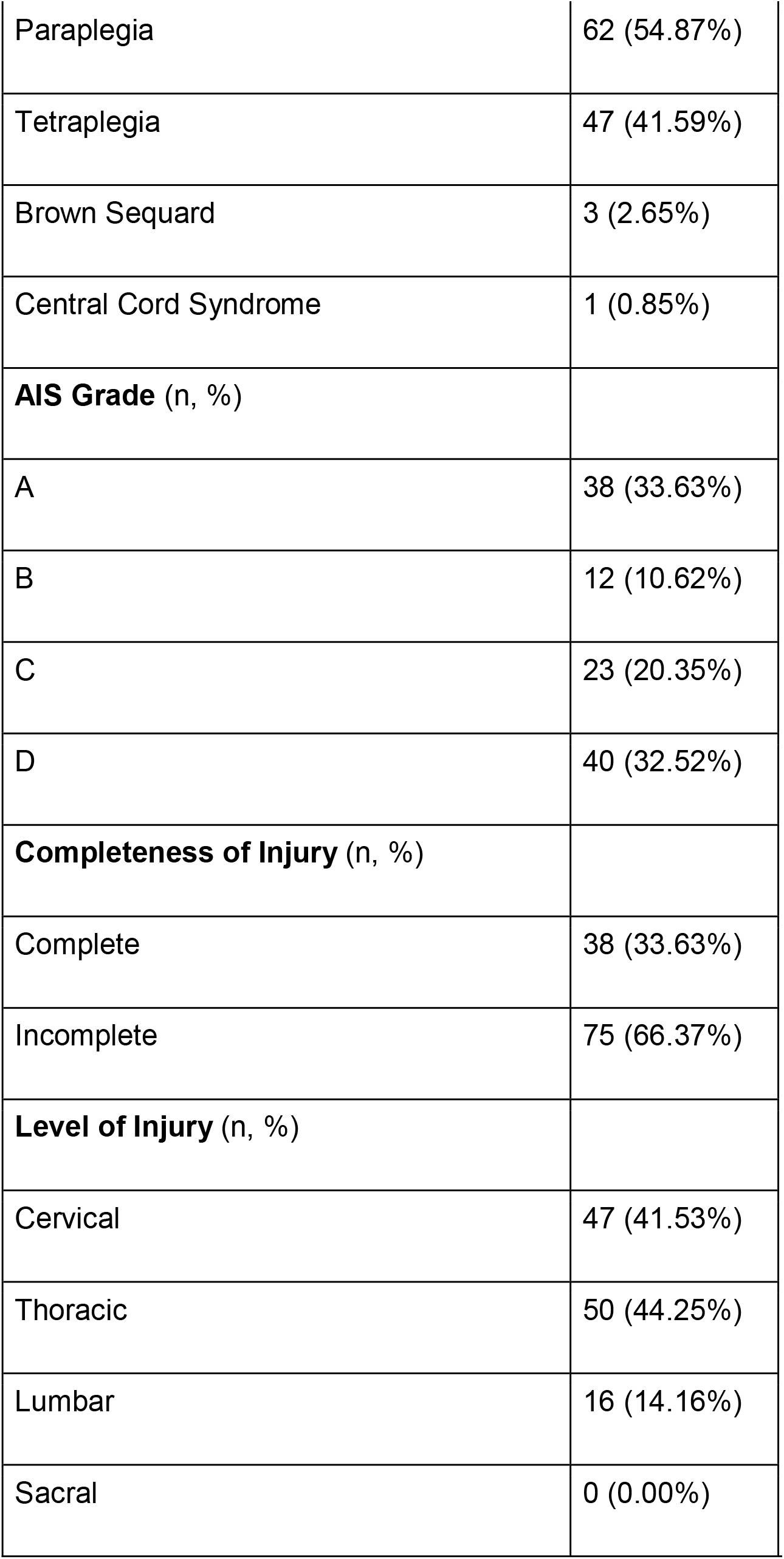
Demographic, lifestyle, and general health data in adults with SCI and uninjured adults.

|  | **Adults with**<br>**SCI**<br>**(n=113)** | **Uninjured community-**<br>**dwelling adults**<br>**(n=471)** |
| --- | --- | --- |
| Age (years, mean ± SD) | 50.52±14.51 | 49.69±17.53 |
| **Sex (n, %)** |  |  |
| Male | 65 (57.52%) | 161 (34.18%) |
| Female | 48 (42.48%) | 309 (65.61%) |
| Other | 0 (0.00%) | 1 (0.21%) |
| **Gender Identity (n, %)** |  |  |
| Male | 65 (57.52%) | 161 (34.18%) |
| Female | 48 (42.48%) | 309 (65.61%) |
| Non-binary | 0 (0.00%) | 0 (0.00%) |
| A-gender | 0 (0.00%) | 1 (0.21%) |
| Other | 0 (0.00%) | 0 (0.00%) |
| **Ethnicity (n, %)** |  |  |
| Hispanic | 12 (10.62%) | 9 (1.91%) |
| Non-Hispanic | 101 (89.38%) | 462 (98.09%) |
| **Racial background** (n, %) |  |  |
| American Indian/Alaskan | 2 (1.77%) | 1 (0.21%) |
| Asian | 2 (1.77%) | 28 (5.94%) |
| Black/African American | 10 (8.85%) | 9 (1.91%) |
| Hawaiian/other Pacific Islander | 1 (0.88%) | 0 (0.00%) |
| Multiracial | 5 (4.42%) | 10 (2.12%) |
| Other | 1 (0.89%) | 6 (1.27%) |
| White | 92 (81.42%) | 417 (88.54%) |
| **Breathing and body awareness** (n,%) |  |  |
| Current body awareness training | 39 (34.51%) | 169 (35.88%) |
| Past body awareness training | 71 (62.83%) | 281 (59.66%) |
| Current breathing exercises | 53 (46.90%) | 195 (41.40%) |
| **Adults with SCI** |  | NA |
| Time after SCI (years, mean ± SD) | 12.84 ± 11.89 | |
| Presence of neuropathic pain (n, %) | 99 (87.61%) |  |
| **Type of SCI** (n, %) |  |  |

|  |  |
| --- | --- |
| Paraplegia | 62 (54.87%) |
| Tetraplegia | 47 (41.59%) |
| Brown Sequard | 3 (2.65%) |
| Central Cord Syndrome | 1 (0.85%) |
| **AIS Grade** (n, %) |  |
| A | 38 (33.63%) |
| B | 12 (10.62%) |
| C | 23 (20.35%) |
| D | 40 (32.52%) |
| **Completeness of Injury** (n, %) |  |
| Complete | 38 (33.63%) |
| Incomplete | 75 (66.37%) |
| **Level of Injury** (n, %) |  |
| Cervical | 47 (41.53%) |
| Thoracic | 50 (44.25%) |
| Lumbar | 16 (14.16%) |
| Sacral | 0 (0.00%) |

### Baseline comparisons of outcomes in adults with SCI vs uninjured adults

At baseline, adults with SCI reported worse interoceptive awareness than uninjured adults on the MAIA-2 dimensions “Not-distracting” [Median(IQR), SCI vs healthy=1.33 (1.17) vs 2.30 (1.50); *p*<.0001] and “Trust” [3.50 (1) vs 3.70 (1.30); *p*=.02] **(Table 2)**.

**Table 2.** Outcome measures in adults with SCI vs uninjured adults at baseline and before and outcome measures in adults with SCI before and after the body awareness interventions.

| **Outcome measures in adults with SCI vs uninjured adults at baseline.** |  |  |  |
| --- | --- | --- | --- |
| **MAIA-2 Dimensions** | **Adults with SCI**<br>**(n=81)** | **Uninjured**<br>**adults**<br>**(n=439)** | **p-value** |
| 1. Noticing (Median, IQR) | 3.50 (1.38) | 3.00 (1.50) | $p=.035^*$ |
| 2. Not Distracting (Median, IQR) | 1.33 (1.17) | 2.30 (1.50) | $p<.0001^*$ |
| 3. Not Worrying (Median, IQR) | 3.00 (0.80) | 3.00 (1.40) | $p=0.83$ |
| 4. Attention Regulation (Median, IQR) | 3.50 (1) | 2.70 (1.30) | $p<.0001^*$ |
| 5. Emotional Awareness (Median, IQR) | 3.75 (1.60) | 3.20 (1.65) | $p=0.11$ |
| 6. Self-Regulation (Median, IQR) | 3.75 (1) | 3.00 (1.5) | $p<.0001^*$ |
| 7. Body Listening (Median, IQR) | 3.75 (1) | 2.30 (1.70) | $p<.0001^*$ |
| 8. Trust (Median, IQR) | 3.50 (1.42) | 3.70 (1.30) | $p=.02^*$ |
| **BARQ-R** | **Adults with SCI**<br>**(n=113)** | **Uninjured**<br>**adults**<br>**(n=471)** | **p-value** |
| BARQ-R (Median, IQR) | 18 (6.5) | 15 (7) | $p<.0001^*$ |

| Outcome measures in adults with SCI before and after the body awareness interventions. |  |  |  |
| --- | --- | --- | --- |
| MAIA-2 Dimension | Adults with SCI<br>(n=43) |  | p-value |
|  | Pre-Intervention | Post-Intervention |  |
| Noticing (Median, IQR) | 3.50 (1.37) | 3.75 (1.50) | $p=.41$ |
| Not-Distracting (Median, IQR) | 1.50 (1.17) | 1.50 (1.34) | $p=.74$ |
| Not-Worrying (Median, IQR) | 3.00 (0.80) | 3.40 (1.40) | $p=.03^*$ |
| Attention Regulation (Median, IQR) | 3.43 (1.32) | 3.43 (1.00) | $p=.09$ |
| Emotional Awareness (Median, IQR) | 3.50 (1.60) | 3.60 (1.65) | $p=.51$ |
| Self-Regulation (Median, IQR) | 3.25 (2.25) | 3.75 (1.25) | $p=.06$ |
| Body Listening (Median, IQR) | 3.00 (2.00) | 3.25 (1.75) | $p=.08$ |
| Trust (Median, IQR) | 3.33 (1.42) | 3.33 (1.25) | $p=.05$ |
| BARQ-R | Adults with SCI<br>(n=69) |  | p-value |
|  | Pre-Intervention | Post-Intervention |  |
| BARQ-R (Median, IQR) | 18 (7) | 16 (7) | $p=.0004^*$ |

*Legend:* BARQ-R: Revised Body Awareness Rating Questionnaire; IQR: Interquartile Range; MAIA-2: Multidimensional Assessment of Interoceptive Awareness, Version 2; SCI: Spinal cord injury; $^*=p<0.05$

However, adults with SCI reported better interoceptive awareness than uninjured adults on MAIA-2 dimensions “Noticing” [3.50 (1.38) vs 3.00 (1.50); *p=*.035], “Attention Regulation” [3.50 (1) vs 2.70 (1.30); *p*<.0001], “Self-regulation” (3.75 (1) vs 3.00 (1.5); *p*<.0001], and “Body Listening” [3.75 (1) vs 2.30 (1.70); *p*<.0001] **(Table 2)**.

Adults with SCI scored higher on BARQ-R, reflecting lower body awareness [18 (6.50) vs 15 (7); *p*<.0001] **(Table 2)**.

### Pre-post intervention comparisons in adults with SCI

Adults with SCI improved on interoceptive awareness after the interventions, represented by a *higher* score on the MAIA-2’s “Not-Worrying” [pre: 3.00 (0.80) vs post: 3.40 (1.40); *p*=.03]. They also improved in body awareness, reflected by a *lower* BARQ-R score [pre: 18 (7) vs post: 16 (7); *p*=.0004] **(Table 2)**.

## Discussion

Adults with SCI demonstrated greater interoceptive awareness than uninjured adults in certain MAIA-2 dimensions, particularly in noticing bodily discomfort vs comfort, changes in breathing, and tuning into body needs. This aligns with Vázquez-Fariñas & Rodríguez-Martin’s (2021) qualitative findings, indicating that adults with SCI often listen for body cues to inform them, e.g., when to use the restroom to catheterize or for their bowel program.^10^

The qualitative feedback received at baseline from our participants with SCI correspond to the results found in the MAIA-2 that many adults with SCI may choose to distract themselves from pain or power through the pain.^4,7^ Extended time spent in a wheelchair often leads to increased body tension, which is also an aspect measured by BARQ-R. Specific interventions which promote present moment awareness and benevolence towards the body lead to less worrying about discomfort or pain, greater ability to ease tension, relax the body, and have greater agency over their pain and life.

## Conclusion

Research on body and interoceptive awareness in adults with SCI is limited. Given the identified deficits in body and interoceptive awareness and the potential for improvement through specific interventions, our results encourage further exploration of how enhancing body and interoceptive awareness can positively impact the quality of their daily lives.

## Data Availability

All data produced in the present study are available upon reasonable request to the authors.

## Statements and Declarations

### Ethical considerations

This study was approved by the University of Minnesota’s Institutional Review Board (IRB) and was conducted in accordance to the Declaration of Helsinki ethical guidelines.

### Consent to participate

All participants gave HIPPA/informed written consent either in person or over UMN secure Zoom before participating in any research activities.

### Consent for publication

Not applicable.

### Data availability

The datasets generated during and/or analyzed during the current study are available from the corresponding author on request.

